# Mapping socioecological interconnections in One Health across human, animal and environmental health: a scoping review protocol

**DOI:** 10.1101/2025.04.02.25325158

**Authors:** Jessica Farias Dantas Medeiros, Leonor Maria Pacheco Santos, Sindy Maciel Silva, Jorge Otávio Maia Barreto, Johnathan Portela da Silva Galdino, Eveline Fernandes Nascimento Vale, Kary Desiree Santos Mercedes, Mayara Suelirta da Costa, Juliana Michelotti Fleck, Karine Suene Mendes Almeida, Verônica Cortez Ginani, Wildo Navegantes de Araújo, Diule Vieira de Queiroz, Christina Pacheco

## Abstract

The One Health framework highlights the interconnectedness of human, animal, and environmental health, requiring interdisciplinary and multisectoral collaboration to address complex global health challenges. This scoping review will systematically map how studies and policy initiatives have incorporated socioecological interconnections within the One Health paradigm (2004 to 2025), following Joanna Briggs Institute guidance and the PRISMA Sc checklist. Searches will be conducted in PubMed, Scopus, Web of Science, LILACS, Health Systems Evidence, Social Systems Evidence, and Google Scholar. The strategy, developed with librarian support and peer reviewed, includes terms in English, Portuguese, and Spanish; pilot searches retrieved 5,333 PubMed and 470 LILACS records. Eligible documents must explicitly present two or more One Health dimensions: food safety, zoonotic diseases, antimicrobial resistance, and environmental health. Data will be extracted using a standardized tool and synthesized in narrative, tabular, and graphical formats. The review will provide a comprehensive mapping of practices and policies, identifying achievements, barriers, and knowledge gaps to inform future strategies and strengthen global health governance.

## 1. Introduction

One Health is a unified, cooperative, transdisciplinary, and interconnected approach to human, animal, and environmental health, aiming to address the challenges posed by emerging and re-emerging diseases, pandemics, zoonoses, antimicrobial resistance, food safety, climate change, and other public health threats [1-3]. Although early ideas linking the environment to health date back to Hayes (1877), Virchow (1855), and Schwabe’s “One Medicine” concept in the 1970s, One Health only consolidated as a formal global strategy in the 21st century [4-7].

A key milestone was the Wildlife Conservation Society’s 2004 report [8], followed by the tripartite alliance between the World Health Organization (WHO), the World Organization for Animal Health (WOAH), and the Food and Agriculture Organization (FAO) in 2010, which recognized shared responsibilities at the human-animal-environment interface [4]. The COVID-19 pandemic demonstrated how the local emergence of a lethal pathogen, likely of animal origin, can rapidly disrupt global public health [9], reinforcing the urgency of coordinated action. In 2022, the worsening climate crisis and the lessons learned from the pandemic led to the formal inclusion of the United Nations Environment Programme (UNEP), strengthening the current quadripartite alliance (WHO, WOAH, FAO, and UNEP), supported by an independent One Health High-Level Expert Panel (OHHLEP) [10].

Furthermore, recent initiatives, such as the Lancet One Health Commission [11], first convened in 2019, emphasized that the socioecological interconnections among humans, animals, and the environment form the foundation of One Health [11]. The Commission noted that while early One Health efforts primarily focused exclusively on zoonotic diseases, recent progress has expanded initiatives but also raised concerns about fragmentation and insufficient conceptual clarity [11]. Another conceptual publication on One Health in 2024 further defined six dimensions of One Health, four of which - control of zoonotic diseases (including epidemics and pandemics from emerging and re-emerging infectious diseases, zoonotic endemics, vector-borne diseases, neglected tropical diseases, and animal-borne diseases), antimicrobial resistance, food safety, and environmental health remain the central focus of attention of One Health worldwide [1].

In this protocol, socioecological interconnection will serve as the guiding criterion for data extraction and narrative synthesis across four dimensions: food safety, zoonoses, antimicrobial resistance, and environmental health. This interconnection will be defined as the simultaneous and interdependent presence of two or more One Health dimensions as reflected in studies, policies, or practices. It encompasses the formulation and implementation of interdisciplinary methodologies, the co-production of evidence, and the explicit consideration of their interdisciplinary impacts and outcomes, including how actions in one dimension affect or depend on others.

Despite institutional and conceptual advances, and the establishment of multiple international One Health networks over the past decade [12], the scientific literature still lacks a review that systematically examines the formulation and implementation of policies and practices addressing socioecological interconnections on a global scale. Preliminary searches in PROSPERO, Cochrane Library, Open Science Framework (OSF), and JBI Evidence Synthesis identified reviews focused on specific aspects: governance (2000–2024) [13], multisectoral collaborations (2018–2024) [14], epidemiological interventions (2024) [15], and regional analyses, such as the One Health triad in public health in Latin America (2016) [16]. Other reviews, such as the bibliometric review published in 2022 [17], highlight data gaps and the persistence of fragmented One Health frameworks. None, however, encompasses the socioecological interconnections of the four core dimensions of One Health from a global, multilingual, and broadly scientific perspective.

In light of this, the present protocol proposes a scoping review to map this conceptual framework, covering scientific evidence and grey literature published between January 2004 and March 27, 2025, in English, Spanish, and Portuguese. The review will examine socioecological interconnections across food safety, zoonoses, antimicrobial resistance, and environmental health in the formulation and implementation of health policies and practices. We aim to analyze how this scenario has evolved over the past 21 years, identify challenges, obstacles, and future priorities. Ultimately, this review will contribute to consolidating perspectives on the global scenario and guide strategies for strengthening the current debate on socioecological interconnections within the One Health framework.

### 1.1. Research questions and aims

The main research questions of this scoping review protocol are the following:

- How have One Health studies addressed the socioecological interconnections of human, animal, and environmental health, particularly across food safety, zoonotic diseases, antimicrobial resistance, and environmental health?
- What do their methodologies, scope, and interdisciplinary collaborations reveal about key achievements, gaps, and future research priorities?

This scoping review protocol aims to map the scientific evidence and gray literature addressing the socioecological interconnections of One Health on a global scale, with emphasis on the food safety, zoonotic diseases, antimicrobial resistance, and environmental health dimensions, to identify advancements, challenges, and future priorities. In order to answer the main objective, the following specific goals were established:

I. Identify and characterize studies, policies, and practices that simultaneously address two or more of the One Health dimensions cited above, analyzing methodologies, scope, and levels of interdisciplinary connection.
II. Examine how these initiatives have operationalized socio-ecological interconnection, including intersectoral coordination, evidence co-production, interdisciplinary methodologies, and reported impacts across dimensions.
III. Systematize the advancements, results, and contributions achieved in implementing the interconnected One Health approach, highlighting experiences in different local, regional, and global contexts.
IV. Identify barriers, challenges, and knowledge gaps that limit the consolidation of the One Health socioecological approach.
V. Propose recommendations for future research, policy formulation, and intersectoral practices, contributing to strengthen interdisciplinary collaboration and address global health threats.

## 2. Materials and Methods

This scoping review will follow the methodological framework developed by the Joanna Briggs Institute (JBI) [18] and the Preferred Reporting Items for Systematic Reviews and Meta-Analyses extension for Scoping Reviews (PRISMA-ScR) checklist [19].

### 2.1. Eligibility Criteria

The mnemonic PCC (Participants, Concept, and Context) was adopted to structure the research question for the scoping review [18]. The PCC for this protocol is defined below and, the reference concepts expanded in Table 1:

**Table 1.** Definition of reference concepts for the scoping review Protocol’s PCC.

|  | Concept | Definition |
| --- | --- | --- |
| Publication of studies, policies, strategies and regulations, addressing the One Health approach | Policy documents | Peer-reviewed scientific studies, as well as institutional frameworks, guidelines, national or international plans, legislation and governance structures, on prevention, promotion, evaluation and management. |
| Socioecological interconnections of One Health, with an emphasis on the dimensions of food safety, zoonotic diseases, antimicrobial resistance and environmental health | Socioecological interconnections of One Health | The simultaneous and interdependent consideration of at least two dimensions of One Health: food safety, zoonotic diseases, antimicrobial resistance, and environmental health. The review will include publications that examine how these interconnections are formulated and implemented in policies, programs, interventions, and practices, as well as how they integrate methodologies, foster interdisciplinary collaboration, and report outcomes or challenges. Special attention will be given to initiatives that demonstrate explicit articulation of cross-sectoral or interdisciplinary impacts |
|  | Food safety | Food safety, avoiding physical, chemical and biological risks throughout the production chain (breeding, slaughter, processing, transportation, marketing) [20]. |
|  | Zoonotic diseases | Zoonoses, or zoonotic diseases, are those transmitted between animals and humans, including those of viral, bacterial, or parasitic origin, which emerge especially in areas where wildlife, domestic animals, and humans interact [21]. Publications regarding zoonotic diseases will be further classified [22] as:<br>EPI: Epidemics and pandemics caused by emerging and reemerging infectious diseases related to the interface between human, animal, plant, and environmental health.<br>END: Zoonotic endemics, vector-borne diseases, neglected tropical diseases, and diseases caused by animals. |
|  | Antimicrobial resistance | Encompasses the emergence, spread and control of resistance to antimicrobials (antibiotics, antivirals, antifungals and antiparasitics) due to the excessive or inappropriate use of these drugs in humans, animals and the environment [23]. |
|  | Environmental health | Refers to the impacts of the environment on human and animal health, such as deforestation, pollution, inadequate waste management, land use, climate change and biodiversity loss [24]. |
| Formulation and implementation of health policies and practices at a global level | Global level | No geographical restrictions will be applied. Publications will be considered across different regions, countries, and settings (local, regional, or international). |

- Participants - Publications of studies, policies, strategies and, regulations, addressing the One Health approach,
- Concept - Socioecological interconnections of One Health, with an emphasis on the dimensions of food safety, zoonotic diseases, antimicrobial resistance and, environmental health,
- Context - Formulation and implementation of health policies and practices at a global level.

Inclusion criteria will follow the PCC framework, incorporating original articles, formally established references, and publications that discuss strategies, experiences, and actions on the socioecological interconnections of One Health, published between 2004 and February 2025, and that directly and substantially address the One Health approach, even if localized.

The protocol also includes gray literature: dissertations and theses, reports from governmental and non-governmental organizations, policy documents, and other relevant sources to ensure a comprehensive overview of the practices and strategies adopted. Letters to the Editor, conference abstracts, event proceedings, sponsored articles, incomplete articles, articles without results, preprints, or articles that were outside the topic of interest of the review will be excluded. An additional search will be conducted in the references of the included and excluded publications.

### 2.2. Search Strategy

The search strategy was developed collaboratively by experts in the concept terms, a librarian, together with other stakeholders. The process included peer review to ensure consistency and methodological rigor. Initially, relevant terms, keywords, and synonyms for the topic were listed in the Medical Subject Headings (MeSH), followed by a pilot search in PubMed, focusing on the concepts of One Health, food safety, zoonotic diseases, antimicrobial resistance, and environmental health. Additional keywords were extracted from titles, abstracts, and indexed terms, ensuring the expansion of the vocabulary of the conceptual block in English, Spanish, and Portuguese (Appendix A, Table A1).

Pilot searches were performed in PubMed and Latin American and Caribbean Literature in Health Sciences (LILACS) via Virtual Health Library (BVS) to validate the combination of descriptors and keywords, employing Boolean operators (“AND”, “OR”) and cross-language synonyms. As presented in Table 2, the PubMed strategy retrieved 5,333 records and the multilingual LILACS strategy retrieved 470 records for the period January 2004 to March 2025. These pilot searches confirmed the adequacy and sensitivity of the conceptual blocks used.

**Table 2.**
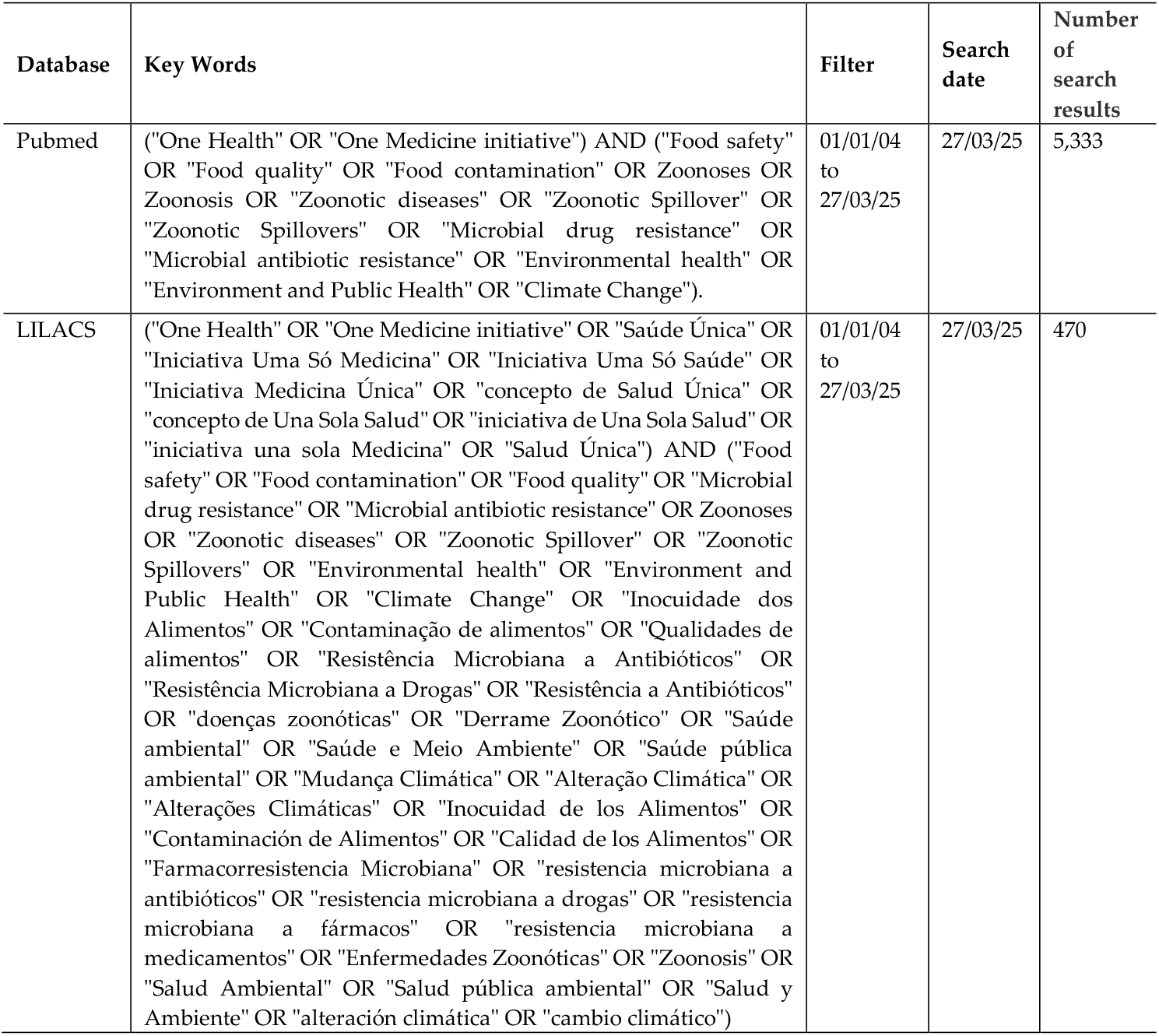
Pilot search in the electronic database PubMed and LILACS.

As evidence sources, we propose the following scientific literature databases: Medical Literature Analysis and Retrieval System Online (MEDLINE) via PubMed (https://pubmed.ncbi.nlm.nih.gov/), Latin American and Caribbean Literature in Health Sciences (LILACS) via Virtual Health Library (BVS) (https://bvsalud.org), Health Systems Evidence (https://www.healthsystemsevidence.org/), Social Systems Evidence (https://www.socialsystemsevidence.org/), SciVerse Scopus (https://www.elsevier.com/products/scopus/search), Web of Science (https://clarivate.com/academia-government/scientific-and-academic-research/research-discovery-and-referencing/web-of-science/). The reference lists of the most relevant studies can also be subject to manual search.

The protocol recommends retrieving grey literature by a structured electronic search on Google Scholar (https://scholar.google.com), in “incognito mode”, thus avoiding the influence of the researcher’s browsing history, examining the first 100 documents retrieved.

### 2.3. Study selection

The studies identified from all the proposed databases will be fed into Mendeley and the Rayyan QCRI software to remove duplicates. Afterward, the studies will be evaluated and selected based on the eligibility criteria by two independent reviewers, following the double-blind methodology when reading the title and summary of the studies in the first phase. In case of disagreements, a third reviewer (master) will be consulted for discussion and final decision. In the second stage, the full text of the selected studies will be read, and those excluded will be justified according to previously established exclusion criteria. Grey literature will be manually included in Mendeley software to manage selected evidence to be cited in the manuscript.

### 2.4. Data Extraction

A data extraction tool was done based on the model provided by JBI (Table 3). The tool includes detailed information about the studies, such as title, authors, year of publication, source, country, publication type, and language. Additionally, data will be collected on the study’s objective, the One Health approach addressed, and methodological characteristics, such as the methodology used, data collection methods, databases consulted, and data collection period.

**Table 3.** Data extraction form.

| Variable | Data to be collected |
| --- | --- |
| Key ID Rayyan | Internal code for study tracking |
| Full reference and web page | Title, year, periodical, volume, issue, pages, authors, web page |
| Publication type | Scientific paper; thesis; dissertation; editorial; protocol; technical document |
| Aims of the study | Objective of the practice or study, as informed by the authors of the article |
| Study context | Documental analysis; field study; laboratory study, as reported by the authors |
| Country and locality of the study | Country and/or locality where the practice or study was carried out, as informed by the authors |
| Data collection period | Data collection period (initial year and final year) reported in the article |
| Primary One Health dimension(s) | Main dimension of the article's object of study:<br>ONE: One Health Policies<br>AMR: Antimicrobial Resistance<br>FSA: Food Safety<br>ENH: Environmental health<br>ZOO/ EPI: Zoonotic epidemics and pandemics, emerging and reemerging infectious diseases caused by animals<br>ZOO/ END: Zoonotic endemics, vector-borne diseases and neglected tropical diseases caused by animals |
| Secondary One Health dimension(s) | Dimension which the article addressed in addition to the main dimension:<br>ONE: One Health Policies<br>AMR: Antimicrobial Resistance<br>FSA: Food Safety<br>ENH: Environmental health<br>ZOO/ EPI: Zoonotic epidemics and pandemics, emerging and reemerging infectious diseases caused by animals<br>ZOO/ END: Zoonotic endemics, vector-borne diseases and neglected tropical diseases caused by animals |
| One Health socioecological interconnections | Direct and substantial presence of two or more dimensions of One Health in joint actions, methodologies, data or decisions, fostering intersectoral and transdisciplinary collaboration (e.g., how actions in one dimension affect or depend on others). |
| Transcript of a relevant direct quote from the One Health socioecological interconnections | Transcription of an excerpt from the interconnected One Health approach |
| Advancements, challenges, and relevant results for the One Health approach | Main results, challenges or advances obtained based on the intervention or practice analyzed |
| Additional information | Additional notes on personal impressions during the extraction process |

### 2.5. Analysis and Presentation of Results

The extracted data will be analyzed and summarized to meet the research objectives and organized into tables, charts, and narrative summaries. The analysis will be based on a narrative synthesis, highlighting the main findings, uncertainties, the evidence’s applicability, strengths, limitations, and implications for managers and future research. Since this is a scoping review, there will be no critical assessment of the methodological quality or analysis of the risk of bias of the included studies [18]. The PRISMA-ScR guideline and the PRISMA Flow Diagram will structure and organize the review [25].

## 3. Conclusions

When applied, the scoping review protocol will be instrumental in revealing several strategies implemented worldwide under the One Health approach, highlighting significant advances in managing food safety, zoonotic diseases, antimicrobial resistance, and environmental health. As a result, challenges related to intersectoral integration and adapting applied policies to local realities may be identified, indicating areas where improvements and new research are needed. We expect to provide a comprehensive understanding of the implementation of the One Health strategy, highlighting both the advances and challenges faced, and to identify practices and that require improvement, contributing to research development, public policies, and the integration of human, animal and environmental health dimensions.

Furthermore, the results of the protocol utilization aim to contribute to formulating future strategies, promoting a more holistic and coordinated approach, improving food safety, tackling zoonotic diseases, combating antimicrobial resistance, and mitigating the impacts of climate change on health. This study also serves as a basis for new research and fosters intersectoral collaboration, essential for the effectiveness of One Health initiatives globally.

## Data Availability

No datasets will be generated or analysed during the current Protocol proposal.

## Author Contributions

**Conceptualization:** JFDM, LMPS, CP, VCG, MSC, WNA, EFNV, JPSG and SMS

**Supervision:** JFDM, JOMB, SMS, LMPS and CP;

**Methodology:** JFDM, JOMB, SMS, CP, LMPS, WNA, VCG, MSC, KDSM, JMF, KSMA, EFNV, JPSG and DVQ

**Software:** JFDM, DVQ, SMS and LMPS

**Validation:** JFDM, JOMB, SMS, CP, LMPS and WNA

**Writing—original draft preparation:** JFDM, SMS, CP, LMPS, VCG and WNA,

**Writing—review and editing:** JFDM, LMPS, CP and WNA

All authors reviewed and approved the final version of this manuscript.

## Funding

This research received no external funding

## Acknowledgments

We are grateful to the collaborators who contributed technical support to the construction of this protocol. We recognize the crucial role of the Translational Research in Public Health discipline’ professors, who provided valuable insights and discussions.

## Conflicts of Interest

The authors declare no conflicts of interest.

## Appendix A

**Table A1.** Conceptual blocks and related terms.

| Conceptual blocks | Related terms |
| --- | --- |
| One Health | One Health<br>One Health concept<br>One Health initiative<br>One Medicine initiative |
| Food safety | Food safety<br>food contamination<br>food quality |
| Zoonotic diseases | Bacterial Zoonoses<br>Viral Zoonoses<br>Fungal zoonoses<br>Protozoan Zoonoses<br>Zoonoses<br>Zoonotic Spillover<br>Zoonotic Spillovers<br>Zoonotic diseases |
| Antimicrobial resistance | Microbial drug resistance<br>Microbial Antibiotic resistance<br>Microbial drug resistance |
| Environmental Health | Environmental Health<br>Health, Environmental<br>Environment and Public Health<br>Climate Change |

**Source:** Prepared by the authors

## Disclaimer/Publisher’s Note

The statements, opinions and data contained in all publications are solely those of the individual author(s) and contributor(s) and not of MDPI and/or the editor(s). MDPI and/or the editor(s) disclaim responsibility for any injury to people or property resulting from any ideas, methods, instructions or products referred to in the content.

## Notes

### Competing Interest Statement

The authors have declared no competing interest.

### Funding Statement

The author(s) received no specific funding for this work.

### Summary of Updates

The Materials and Methods section was revised.

